# Deep Learning-Based Risk Prediction Model for Major Adverse Cardiovascular Events in Long-Term Breast Cancer Survivors

**DOI:** 10.1101/2025.06.16.25329733

**Authors:** Si Nae Oh, Jae Yong Shim, Yong Je Lee, Ji Won Lee

## Abstract

**Background:** Clinical practice guidelines recommend cardiovascular toxicity risk restratification including evaluation of new cardiovascular risk factors and cardiovascular diseases 5 years after cancer therapy in adult cancer survivors. We aimed to develop prediction models for the risk of 10-year cardiovascular complications in five-year breast cancer survivors.

**Methods:** We used the Korean National Health Insurance Service databases between 2005 and 2021, including 5,131 five-year female breast cancer survivors diagnosed in 2006. The study population was randomly split in a 4:1 ratio into the derivation and validation cohort. The primary outcome was the occurrence of major adverse cardiovascular events (MACEs) at any time before the final follow-up at 10 years. We developed a deep learning survival model (DeepSurv) and compared its performance with traditional Cox proportional hazards (CPH) model using the same dataset. Model performance was assessed by time-dependent concordance (C^td^) index. Shapley Additive Explanations (SHAP) was used to assess feature importance.

**Results:** In the validation cohort, the DeepSurv and CPH model yielded C^td^ index values of 0.738 (95% CI, 0.712–0.779) and 0.733 (95% CI, 0.649–0.777), respectively. In SHAP analysis, age, history of stroke, dyslipidemia, anthracycline use, and aromatase inhibitor therapy ranked highly in both models.

**Conclusions:** A deep learning survival model that incorporates both conventional and breast cancer treatment-related cardiovascular risk factors outperformed traditional regression model in predicting 10-year MACEs among individual five-year breast cancer survivors.

## INTRODUCTION

Five-year breast cancer survival rates now exceed 90% in most high-income countries, contributing to a global population of over 7.7 million breast cancer survivors (1,2). These survivors have a significantly increased risk of cardiovascular disease (CVD) and cardiovascular mortality, due to shared risk factors underlying both cancer and CVD (3). In addition, commonly used breast cancer treatments, such as chemotherapy, radiation therapy, and biologic agents, can cause long-term cardiac complications that may manifest throughout a patient’s lifetime (3).

While prior research and clinical attention have largely focused on cardiotoxicity occurring during or early after cancer treatment, less emphasis has been placed on the prediction and prevention of late-onset cardiovascular complications in this population (4). Recent clinical practice guidelines recommend reassessing cardiovascular toxicity risk five years after cancer therapy in asymptomatic adult cancer survivors (5,6). These guidelines have provided risk stratification criteria based on both conventional and cancer treatment related cardiovascular risk factors, including the evaluation of newly developed cardiovascular risk factors and cardiovascular diseases (5,6). Long-term follow-up surveillance should be organized and integrated into the long-term cancer survivorship care based on this stratification (5, 6). This includes patient education and cardiovascular risk factor optimization, in collaboration with primary care providers or specialists with expertise in cardiovascular risk factor management (5). For high-risk survivors, regular monitoring with complementary tests, such as electrocardiography, natriuretic peptides measurement, and echocardiography is recommended (5).

However, the diagnostic value of these stratification criteria has not yet been studied in adult cancer survivors, in contrast to the extensively studied guidelines for adult survivors of childhood cancer (7–9). Furthermore, while current criteria include anthracycline use and radiation therapy as treatment-related risk factors, breast cancer treatment regimens frequently also include HER2-targeted therapies and endocrine treatments, which may influence long-term cardiovascular risk in this population (10,11,12). Therefore, there is an urgent need for prediction models to estimate individual long-term cardiovascular risk in breast cancer survivors, enabling individualized surveillance and prevention strategies (4).

The objective of our study was to develop and validate a 10-year risk prediction models for major adverse cardiovascular events (MACEs) in individual 5-year breast cancer survivors using the Korean National Health Insurance Service (NHIS) databases. Using deep learning survival model, both conventional and breast cancer treatment-related cardiovascular risk factors were integrated into a prediction model.

## METHODS

### Ethical approval

This study was approved by the Institutional Review Board of National Health Insurance Service Ilsan Hospital (IRB number NHIMC 2021-11-001) and the requirement for informed consent was waived for the NHIS database is anonymized.

### Data sources

We utilized data from the National Health Insurance Service (NHIS) databases spanning 2005 to 2021. The NHIS, South Korea’s single-payer non-profit health insurer, provides universal coverage and maintains a comprehensive electronic database that includes individual-level information on inpatient and outpatient claims, prescriptions, diagnoses, procedures, and treatments. De-identified, customized datasets can be made available to researchers upon request (13). Additionally, we incorporated data from the National Health Screening Program, which offers biennial mandatory health examinations to all NHIS beneficiaries aged 40 years and older. This screening includes self-reported lifestyle and health behavior questionnaires, anthropometric data, blood pressure measurements, and laboratory tests (14). The validity of the NHIS database has been documented elsewhere (14).

### Study design and population

This nationwide population-based retrospective cohort study included female breast cancer survivors who were newly diagnosed with breast cancer in 2006 and survived for at least five years, until 2011. We identified 14,170 patients who were newly diagnosed with breast cancer in 2006. We excluded 64 male patients, 1,984 patients who had missing age data, 6,774 patients without health screening records or not subject to health screening, and 217 patients died before the index date (January 1, 2012). The final cohort of 5,131 participants was randomly divided in a 4:1 ratio to derivation and validation sets. During the training process, 20% of the derivation cohort was held out for internal evaluation, and final model performance was assessed in the validation cohort. The cohort selection process is illustrated in Figure 1.

**Figure 1.**
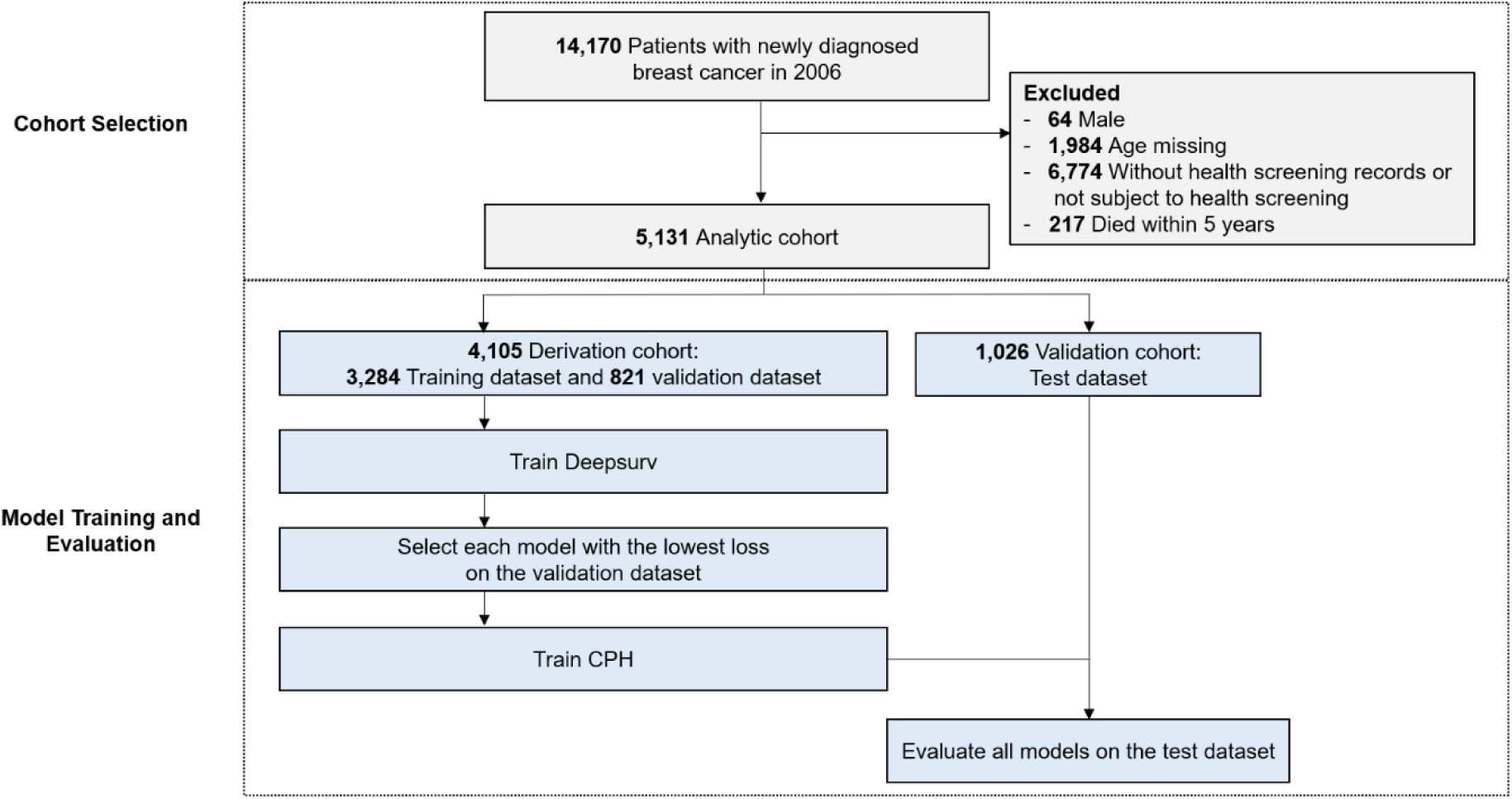
Flow chart of cohort selection and model training and evaluation. CPH, the Cox proportional hazards model.

### Major adverse cardiovascular events

The primary outcome was the occurrence of MACE (a composite of acute myocardial infarction, congestive heart failure, stroke, and all-cause death) at any time before the final follow-up at 10 years (December 31, 2021) (15). Acute myocardial infarction was defined as hospitalization for primary or secondary diagnosis of the International Classification of Diseases, Tenth Revision (ICD-10) codes I21 and I22. Patients were considered to have congestive heart failure if they were hospitalized for primary or secondary diagnosis of ICD-10 codes I50. A stroke was defined as hospitalization for primary or secondary diagnosis of ICD-10 codes I60 to I69. The ICD-10 codes were derived from the American Heart Association guidelines (16).

### Data collection

To construct the models, we collected 26 clinical variables including socio-demographic characteristics, lifestyle behavior, physical examination and laboratory tests, comorbidities, prior cancer treatment, and prior cardiovascular disease. A detailed list of the variables included and tested is provided in Table 1. Socio-demographic characteristics included age (years) and household income (upper half and lower half) at index date. Household income was derived from insurance premiums. Lifestyle behavior included cigarette smoking (non-smoker and smoker), alcohol consumption (non-drinker and drinker), and physical activity (none and 1 day per week or more) within 2 years of index date. Physical examination and laboratory tests included body mass index (kg/m2), systolic blood pressure (mmHg), diastolic blood pressure (mmHg), fasting serum glucose (mg/dL), total cholesterol (mg/dL), creatinine (mg/dL), and hemoglobin (g/dL) within 2 years of index date. The body mass index was calculated by dividing the participant’s weight in kilograms by the square of their height in meters. Comorbidities included hypertension, diabetes mellitus, and dyslipidemia, and chronic kidney disease within 2 years of index date.

**Table 1.** Baseline characteristics.

|  | **Total**<br>**(N = 5,131)** | **Derivation**<br>**cohort (N =**<br>**4,105)** | **Validation**<br>**cohort (N =**<br>**1,026)** | **P-value** |
| --- | --- | --- | --- | --- |
| Age, y, mean (SD) | 56.2 (9.5) | 56.2 (9.5) | 56.2 (9.7) | 0.819 |
| Household income |  |  |  |  |
| Lower half | 1,879 (36.6) | 1,489 (36.3) | 390 (38.0) | 0.301 |
| Upper half | 3,252 (63.4) | 2,616 (63.7) | 636 (62.0) |  |
| Prior cancer treatment |  |  |  |  |
| Anthracycline | 2,056 (40.1) | 1,654 (40.3) | 402 (39.2) | 0.516 |
| Trastuzumab | 38 (0.7) | 29 (0.7) | 9 (0.9) | 0.568 |
| Endocrine therapy |  |  |  |  |
| Tamoxifen | 2,045 (39.9) | 1,636 (39.9) | 406 (39.9) | 0.996 |
| Aromatase inhibitors | 1,333 (26.0) | 1,068 (26.0) | 265 (25.8) | 0.902 |
| Radiotherapy | 2,419 (47.1) | 1,949 (47.5) | 470 (45.8) | 0.338 |
| Prior cardiovascular disease |  |  |  |  |
| Myocardial infarction | 115 (2.2) | 93 (2.3) | 22 (2.1) | 0.814 |
| Stroke | 636 (12.4) | 509 (12.4) | 127 (12.4) | 0.985 |
| Congestive heart failure | 173 (3.4) | 136 (3.3) | 37 (3.6) | 0.642 |
| Peripheral artery occlusive disease | 805 (15.7) | 635 (15.5) | 170 (16.6) | 0.386 |
| Atrial fibrillation | 135 (2.6) | 107 (2.6) | 28 (2.7) | 0.827 |
| Comorbidities |  |  |  |  |
| Hypertension | 1,962 (38.2) | 1,580 (38.5) | 382 (37.2) | 0.458 |
| Diabetes mellitus | 729 (14.2) | 572 (13.9) | 157 (15.3) | 0.262 |
| Dyslipidemia | 2,143 (41.8) | 1,702 (41.5) | 441 (43.0) | 0.377 |
| Chronic kidney disease | 356 (6.9) | 298 (7.3) | 58 (5.7) | 0.070 |
| Cigarette smoking |  |  |  |  |
| Non-smoker | 5,060 (98.6) | 4,054 (98.8) | 1,006 (98.1) | 0.083 |
| Smoker | 71 (1.4) | 51 (1.2) | 20 (2.0) |  |
| Alcohol consumption |  |  |  |  |
| Non-drinker | 4,587 (89.4) | 3,664 (89.3) | 923 (90.0) | 0.740 |
| 1 days/week | 374 (7.3) | 300 (7.3) | 74 (7.2) |  |
| 2 days/week | 96 (1.9) | 81 (2.0) | 15 (1.5) |  |
| ≥3 days/week | 74 (1.4) | 60 (1.5) | 14 (1.4) |  |
| Moderate-to-vigorous Physical activity |  |  |  |  |
| None | 2,259 (44.0) | 1,827 (44.5) | 432 (42.1) | 0.089 |
| 1 day/week | 273 (5.3) | 230 (5.6) | 43 (4.2) |  |
| 2 days/week | 528 (10.3) | 416 (10.1) | 112 (10.9) |  |
| ≥3 days/week | 2,071 (40.4) | 1,632 (40.0) | 439 (42.8) |  |
| BMI, kg/m<sup>2</sup>, mean (SD) | 23.6 (3.1) | 23.6 (3.1) | 23.4 (3.0) | 0.129 |
| Systolic blood pressure, mmHg, mean (SD) | 121.3 (15.7) | 121.2 (15.8) | 121.8 (15.7) | 0.295 |
| Diastolic blood pressure, mmHg, mean (SD) | 75.1 (10.1) | 75.0 (10.2) | 75.5 (9.9) | 0.308 |
| Fasting serum glucose, mg/dl, mean (SD) | 97.9 (21.3) | 97.6 (20.5) | 99.1 (24.3) | 0.046 |
| Total cholesterol, mg/dL, mean (SD) | 197.9 (37.4) | 197.9 (37.7) | 197.8 (36.4) | 0.964 |
| Creatinine, mg/dL, mean (SD) | 0.9 (0.7) | 0.9 (0.7) | 0.9 (0.7) | 0.882 |
| Hemoglobin, g/dL, mean (SD) | 12.8 (1.1) | 12.8 (1.1) | 12.9 (1.1) | 0.153 |
| MACE during follow-up period | 714 (13.9) | 573 (14.0) | 141 (13.7) | 0.858 |
| Myocardial infarction | 26 (0.5) | 21 (0.5) | 5 (0.5) | 0.922 |
| Stroke | 254 (5.0) | 202 (4.9) | 52 (5.1) | 0.846 |
| Congestive heart failure | 60 (1.2) | 51 (1.2) | 9 (0.9) | 0.330 |
| All-cause death | 453 (8.8) | 363 (8.8) | 90 (8.8) | 0.943 |

All values are presented as frequency (%) unless otherwise specified.
BMI, body mass index; MACE, major adverse cardiovascular event; and SD, standard deviation.

### Model training and performance evaluation

We developed a model based on deep learning survival analysis (DeepSurv) as shown in figure 2 (17). To compare its performance against traditional methods, we also trained a Cox proportional hazard regression (CPH) model using the same data set. DeepSurv model was implemented using the Python module Pycox (version 0.2.3), while the CPH model was implemented with scikit-survival (version 0.21.0). DeepSurv hyperparameters were tuned with Optuna (version 3.5.0) using five-fold cross-validation.

**Figure 2.**
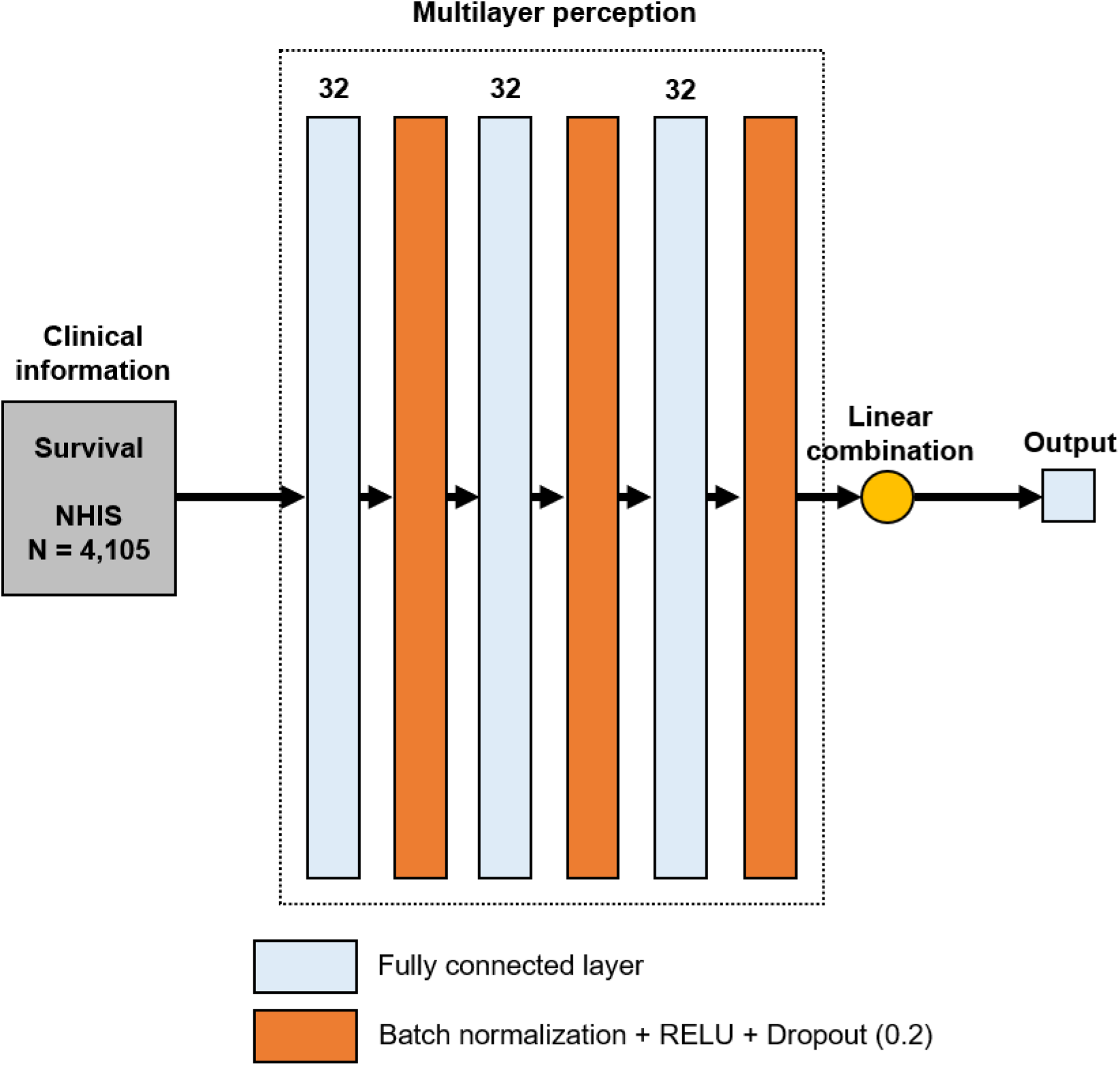
Deep learning model architecture. Clinical information was input into the DeepSurv model, a multilayer perceptron architecture designed to predict an individual’s risk of major adverse cardiovascular events (MACEs). The model outputs a single node representing the patient’s log risk score, which is subsequently used to parameterize the Weibull survival distribution and compute the weight *w*, corresponding to the individualized survival probability. To improve the model performance and prevent overfitting, we applied weight decay regularization, RELU activation with batch normalization, dropout, and Adaptive Moment estimation (Adam) optimization. NHIS, National Health Insurance Service database; and RELU, Rectified Linear Unit.

For model evaluation, we used the time-dependent concordance (C^td^) index (18), an extension of Harrell’s concordance index (C index) (19), which is the most widely used metric for assessing discrimination performance in survival analysis. A C^td^ index value of 0.5 reflects random prediction, while a value of 1 indicates perfect prediction. To estimate 95% confidence intervals for these metrics, bootstrap resampling with 1,000 iterations was employed. The model training and evaluation workflow is depicted in Figure 1. We also used Shapley Additive exPlanations (SHAP) to quantify the influence of individual features on model predictions using the SHAP Python module (version 0.43.0). (20)

Data processing was conducted using SAS (version 9.4), while all analysis codes were implemented and performed in Python (version 3.8). Data analysis was performed from 1 June 2023 to 31 December 2024.

## RESULTS

### Cohort characteristics

The study included a total of 5,131 patients who were divided in a 4:1 ratio to either the derivation or validation cohorts. Baseline characteristics of the cohort are presented in Table 1. The mean (SD) population age was 56.2 (9.5) years. 40.1% of the patients received chemotherapy with a regimen containing anthracycline and 0.7% received treatment with antihuman epidermal growth factor receptor antibodies. 39.9% of the patients were treated with tamoxifen and 26.0% received aromatase inhibitors. 47.1% of the patients underwent radiotherapy. During the 48,054 person-years of follow-up, 714 patients developed MACE. This corresponds to incidence rates of 14.9 per 1,000 person-years.

### Predictive performance evaluation

We trained the Deepsurv and traditional CPH models using the derivation cohort and evaluated their performance in the validation cohort. To assess the performance of the models, we calculated the C^td^ index. The C^td^ index values for both models in the derivation and validation cohorts are shown in Table 2. In validation cohort, the CPH model yielded C^td^ index of 0.733 (95% CI 0.649–0.777), while the DeepSurv model demonstrated slightly better performance with a C^td^ index of 0.738 (95% CI 0.712–0.779).

**Table 2.** Performance comparison of deep learning and traditional model for 10-year cardiovascular disease risk prediction among 5-year breast cancer survivors.

| | $C^{td}$ (95% CI) | |
| --- | --- | --- |
|  | Cox proportional hazards model | DeepSurv |
| Derivation cohort | 0.738 (0.712–0.761) | 0.752 (0.737–0.769) |
| Validation cohort | 0.733 (0.649–0.777) | 0.738 (0.712–0.779) |

CI, confidence interval; and $C^{td}$ , the time-dependent concordance index.

SHAP values were used to identify which clinical factors had the greatest contributions to the prediction model. Age, history of stroke, dyslipidemia, anthracycline use, aromatase inhibitor therapy consistently ranked among the most important predictors in both the DeepSurv and CPH models (Figure 3 and Figure S1).

**Figure 3.**
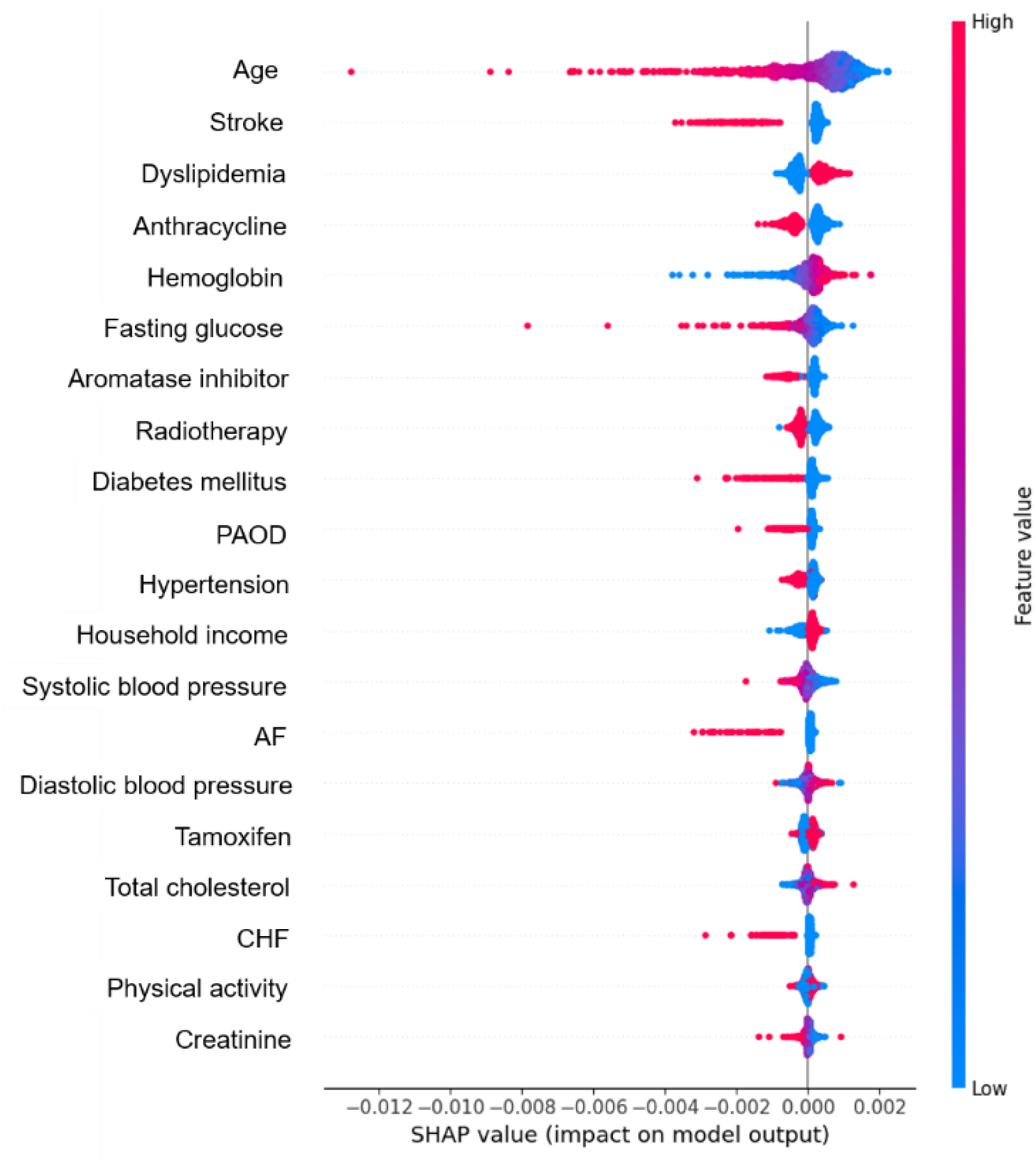
Shapley Additive Explanations (SHAP) values of Deepsurv model. The summary plot displays the impact of individual features on the model’s predictions, demonstrating the relationship between feature values and their estimated impact on major adverse cardiovascular events (MACEs). Predictor importance decreases from top to bottom along the y-axis. Negative SHAP values indicate an increased predicted risk of MACE, whereas positive values indicate a decreased predicted risk. Binary variables are encoded as Low for No, High for Yes. AF, atrial fibrillation; CHF, congestive heart failure; and PAOD, peripheral arterial obstructive disease.

## DISCUSSION

We developed and validated a prediction model for 10-year MACEs risk in 5-year breast cancer survivors, using large population-based cohort. We implemented deep learning survival model to integrate breast cancer therapy-related and conventional cardiovascular risk factors into a prediction model and demonstrated its superiority to the traditional model in predicting chronic cardiovascular complications.

### Conventional cardiovascular risk factors in patients with BC

Recent prediction models for early cardiotoxicity risk among patients with breast cancer have shown good ability to predict MACE, based on conventional cardiovascular risk factors and/or current cancer treatment (21,22). For long-term cardiovascular surveillance in adult cancer survivors, clinical practice guidelines have suggested risk stratification criteria based on both conventional and cancer treatment related cardiovascular risk factors (5,6). These conventional cardiovascular risk factors include medical risk factors (hypertension, diabetes, dyslipidemia, chronic kidney disease) and lifestyle risk factors (smoking, alcohol intake, exercise, obesity).

Our findings from Shapley Additive Explanations regarding traditional cardiovascular risk factors align with previous literature. However, in our study, presence of dyslipidemia was associated with a lower risk of MACE. We defined dyslipidemia cases as individuals prescribed antidyslipidemic medications under ICD-10 code for dyslipidemia. Statins, an essential approach for current lipid-lowering therapies, are well known to improve cardiovascular outcomes. A recent observational study reported that statin therapy was associated with a reduced risk of MACE in patients with breast cancer undergoing breast-conserving surgery and adjuvant whole breast radiotherapy (23). Further research is warranted to explore the potential role of lipid-lowering therapy in mitigating cardiovascular complications in this population.

Our results also showed that lower diastolic blood pressure was associated with an increased risk of MACE. While high diastolic blood pressure is associated with vascular and organ damage in individuals with preserved vascular compliance, the relationship between diastolic blood pressure and cardiovascular risk becomes more complex in cases of noncompliant vasculature, displaying a U-shaped association (24). Multiple studies in patients with heart failure with preserved ejection fraction have reported that low diastolic blood pressure, particularly when combined with high systolic blood pressure, is linked to myocardial damage, coronary heart disease, heart failure hospitalization, stroke, and cardiovascular mortality (25,26,27).

### BC therapy-related cardiovascular risk factors

It is well established that anthracycline chemotherapy and HER2-targeted therapies trigger cardiac dysfunction (28). Anthracycline-induced cardiotoxicity can occur in different phases, including acute toxicity (within days), early-onset chronic toxicity (within the first year), and late-onset chronic toxicity (developing years to decades after treatment) (29). The majority of cardiotoxic events occur within the first year following anthracycline exposure, while the long-term effects beyond 10 years in breast cancer survivors remain poorly characterized (29). In our study, anthracycline was a significant risk factor for MACE among long-term breast cancer survivors.

In contrast, trastuzumab was not a significant risk factor for MACE in our analysis. There could be several explanations for the inconsistent results. The incidence of trastuzumab-related symptomatic heart failure is relatively low (0-4%) (30,31). Also, in contrast to anthracycline-induced cardiotoxicity, trastuzumab exposure can result in cardiac dysfunction and heart failure that is mostly reversible (32). Third, since we included patients diagnosed with breast cancer in 2006, only 0.7% of the patients in this study administered trastuzumab. Therefore, our findings may not be generalized, and careful monitoring of CVDs is essential for long-term breast cancer survivors who have received anthracycline and HER2-targeted therapies (6).

Endocrine therapy is widely used, as approximately 65–70% of patients with early or metastatic breast cancer have hormone receptor–positive disease (33). Adjuvant treatment with tamoxifen or aromatase inhibitors in early-stage hormone receptor–positive breast cancer has been shown to lower recurrence rates and enhance overall survival (33). Tamoxifen is the endocrine therapy of choice for premenopausal women, whereas strategies in postmenopausal women can include tamoxifen, aromatase inhibitors, or a sequential combination (33). In the adjuvant setting, endocrine therapy is typically administered over an extended duration, often for five years or longer, underscoring the need for a detailed evaluation of its long-term toxicity (3). A meta-analysis reported that long-term aromatase inhibitors use was associated with an increased risk for hypercholesterolemia and CVD compared with tamoxifen use (11,12). Meanwhile, although tamoxifen positively impacts lipid profiles, it has not demonstrated a protective effect on cardiovascular outcomes and has been shown to minimally increase the risk of venous thromboembolism compared to the use of aromatase inhibitors (12). In our study, both tamoxifen and aromatase inhibitors were associated with increased risk of MACE.

Additionally, we found that radiation therapy was associated with an elevated risk of MACE. Previous studies reported that there was an excess of non–breast cancer deaths after 5 years among patients receiving radiation therapy, mainly due to CVD and lung cancer (34). Radiation therapy that includes the heart within the treatment field carry the risk of long-term coronary artery disease and heart failure, which can manifest as early as 5 years after exposure, with the risk persisting for up to 30 years (35,36). Despite introduction of heart-sparing radiation techniques, incidental heart irradiation, even of smaller volumes, is associated with cardiac perfusion defects, indicating an association between the mean heart radiation dose and the incidence of radiation-induced coronary heart disease, without a clear threshold for safety (37,38).

### Deep learning in risk prediction modeling

By using CPH and Deepsurv to develop risk prediction model, we compared traditional statistical approaches with a novel machine-learning-based method in survival analysis. We demonstrated that the Deepsurv models outperformed conventional models in predicting future cardiovascular outcomes for breast cancer survivors. Deep learning network models, such as DeepSurv, can improve the performance by effectively capturing complex and non-linear interactions among variables (17). However, our results also indicate that traditional models currently in use are capable of accurately predicting cardiovascular events, suggesting that the contribution of non-linear interactions among the predictors in our study might be limited.

### Limitations

This study has several potential limitations. First, the use of HER-2 targeted therapy was low in our cohort, as only 0.7% of patients received trastuzumab due to the study’s inclusion of patients diagnosed in 2006, which may limit the generalizability of our findings to contemporary breast cancer populations. Second, due to our reliance on administrative data, we were unable to account for certain risk factors like laterality of radiation therapy, family history of CVD, echocardiographic parameters, electrocardiography, blood biomarkers, and genetic variants. Incorporating these parameters could potentially enhance the model’s performance. Lastly, our prediction model requires prospective external validation before recommending its use as a clinical decision support tool to personalize cardiovascular surveillance and preventive strategies.

## CONCLUSION

We developed and validated a risk prediction model to estimate the 10-year cardiovascular risk in long-term breast cancer survivors, using nationwide population-based cohort. This study showed that deep learning model, incorporating both patient-related and cancer treatment-related risk factors, could improve the prediction that outperform traditional models. Further research will enable us to refine this prediction system further, aiming for greater accuracy and customization to tailor long-term cancer survivorship programs to individual cardiovascular risk.

## Data Availability

The data that support the findings of this study are available from National Health Insurance Service but restrictions apply to the availability of these data, which were used under license for the current study, and so are not publicly available. Data are however available from the corresponding author upon reasonable request and with permission of National Health Insurance Service.

## Abbreviations and Acronyms

CPH: the Cox proportional hazards model
CVD: cardiovascular disease
MACE: major adverse cardiovascular events
NHIS: the Korean National Health Insurance Service

## Acknowledgement

This study used the National Health Insurance Service database (NHIS-2022-1-411). The interpretations and conclusions reported herein do not represent those of the National Health Insurance Service.

## References

1. Bray F, Laversanne M, Sung H, et al. Global cancer statistics 2022: GLOBOCAN estimates of incidence and mortality worldwide for 36 cancers in 185 countries. CA Cancer J Clin 2024;74:229–263.

2. Giaquinto AN, Sung H, Newman LA et al. Breast cancer statistics 2024. CA Cancer J Clin 2024;74:477–495.

3. Mehta LS, Watson KE, Barac A, et al. Cardiovascular Disease and Breast Cancer: Where These Entities Intersect: A Scientific Statement From the American Heart Association. Circulation. 2018;137(8):e30–e66.

4. Kaboré EG, Macdonald C, Kaboré A, et al. Risk Prediction Models for Cardiotoxicity of Chemotherapy Among Patients With Breast Cancer: A Systematic Review. JAMA Netw Open. 2023;6(2):e230569.

5. Lyon AR, López-Fernández T, Couch LS, et al. 2022 ESC Guidelines on cardio-oncology developed in collaboration with the European Hematology Association (EHA), the European Society for Therapeutic Radiology and Oncology (ESTRO) and the International Cardio-Oncology Society (IC-OS). Eur Heart J. 2022;43(41):4229–4361.

6. Armenian SH, Lacchetti C, Barac A, et al. Prevention and Monitoring of Cardiac Dysfunction in Survivors of Adult Cancers: American Society of Clinical Oncology Clinical Practice Guideline. J Clin Oncol. 2017;35(8):893–911.

7. Leerink JM, de Baat EC, Feijen EAM, et al. Cardiac Disease in Childhood Cancer Survivors: Risk Prediction, Prevention, and Surveillance: JACC CardioOncology State-of-the-Art Review. JACC CardioOncol. 2020;2(3):363–378.

8. Chaix MA, Parmar N, Kinnear C, et al. Machine Learning Identifies Clinical and Genetic Factors Associated With Anthracycline Cardiotoxicity in Pediatric Cancer Survivors. JACC CardioOncol. 2020;2(5):690–706.

9. Ehrhardt MJ, Liu Q, Mulrooney DA, et al. Improved Cardiomyopathy Risk Prediction Using Global Longitudinal Strain and N-Terminal-Pro-B-Type Natriuretic Peptide in Survivors of Childhood Cancer Exposed to Cardiotoxic Therapy. J Clin Oncol. 2024. 10.1200/jco.23.01796.Jco2301796.

10. Tan-Chiu E, Yothers G, Romond E et al. Assessment of cardiac dysfunction in a randomized trial comparing doxorubicin and cyclophosphamide followed by paclitaxel, with or without trastuzumab as adjuvant therapy in node-positive, human epidermal growth factor receptor 2-overexpressing breast cancer: NSABP B-31. J Clin Oncol 2005;23:7811–9.

11. Amir E, Seruga B, Niraula S, Carlsson L, Ocaña A. Toxicity of adjuvant endocrine therapy in postmenopausal breast cancer patients: a systematic review and meta-analysis. J Natl Cancer Inst. 2011;103(17):1299–1309.

12. Yu Q, Xu Y, Yu E, Zheng Z. Risk of cardiovascular disease in breast cancer patients receiving aromatase inhibitors vs. tamoxifen: A systematic review and meta-analysis. J Clin Pharm Ther. 2022;47(5):575–587.

13. Kim HK, Song SO, Noh J, Jeong IK, Lee BW. Data Configuration and Publication Trends for the Korean National Health Insurance and Health Insurance Review & Assessment Database. Diabetes Metab J. 2020;44(5):671–678.

14. Seong SC, Kim YY, Park SK, et al. Cohort profile: the National Health Insurance Service-National Health Screening Cohort (NHIS-HEALS) in Korea. BMJ Open. 2017;7(9):e016640.

15. Bosco E, Hsueh L, McConeghy KW, Gravenstein S, Saade E. Major adverse cardiovascular event definitions used in observational analysis of administrative databases: a systematic review. BMC Med Res Methodol. 2021;21(1):241.

16. Virani SS, Alonso A, Aparicio HJ, et al. Heart Disease and Stroke Statistics-2021 Update: A Report From the American Heart Association. Circulation. 2021;143(8):e254–e743.

17. Katzman JL, Shaham U, Cloninger A, Bates J, Jiang T, Kluger Y. DeepSurv: personalized treatment recommender system using a Cox proportional hazards deep neural network. BMC Med Res Methodol 2018;18:24.

18. Antolini L, Boracchi P, Biganzoli E. A time-dependent discrimination index for survival data. Stat Med. 2005;24(24):3927–3944.

19. Harrell FE, Jr., Lee KL, Mark DB. Multivariable prognostic models: issues in developing models, evaluating assumptions and adequacy, and measuring and reducing errors. Stat Med. 1996;15(4):361–387.

20. Lundberg SM, Lee S. A unified approach to interpreting model predictions. Adv. Neural Inform Process Syst. 2017; 30:4765–4774.

21. Abdel-Qadir H, Thavendiranathan P, Austin PC, et al. Development and validation of a multivariable prediction model for major adverse cardiovascular events after early stage breast cancer: a population-based cohort study. Eur Heart J. 2019;40(48):3913–3920.

22. Kim DY, Park MS, Youn JC, et al. Development and Validation of a Risk Score Model for Predicting the Cardiovascular Outcomes After Breast Cancer Therapy: The CHEMO-RADIAT Score. J Am Heart Assoc. 2021;10(16):e021931.

23. Huang YJ, Lin JA, Chen WM, Shia BC, Wu SY. Statin Therapy Reduces Radiation-Induced Cardiotoxicity in Patients With Breast Cancer Receiving Adjuvant Radiotherapy. J Am Heart Assoc 2024;13:e036411.

24. Lee H, Yano Y, Cho SMJ, Park S, Lloyd-Jones DM, Kim HC. Cardiovascular Risk of Isolated Diastolic Hypertension Defined by the 2017 American College of Cardiology/American Heart Association Blood Pressure Guideline: A Nationwide Age-Stratified Cohort Study. Hypertension. Dec 2020;76(6):e44–e46. doi:10.1161/hypertensionaha.120.16018

25. McEvoy JW, Chen Y, Rawlings A, et al. Diastolic Blood Pressure, Subclinical Myocardial Damage, and Cardiac Events: Implications for Blood Pressure Control. J Am Coll Cardiol. Oct 18 2016;68(16):1713–1722. doi:10.1016/j.jacc.2016.07.754

26. Suzuki K, Claggett B, Minamisawa M, et al. Pulse Pressure, Prognosis, and Influence of Sacubitril/Valsartan in Heart Failure With Preserved Ejection Fraction. Hypertension. Feb 2021;77(2):546–556. doi:10.1161/hypertensionaha.120.16277

27. Sandesara PB, O’Neal WT, Kelli HM, Topel M, Samman-Tahhan A, Sperling LS. Diastolic Blood Pressure and Adverse Outcomes in the TOPCAT (Treatment of Preserved Cardiac Function Heart Failure With an Aldosterone Antagonist) Trial. J Am Heart Assoc. Feb 23 2018;7(5)doi:10.1161/jaha.117.007475

28. Dauccia C, Agostinetto E, Arecco L et al. Cardiovascular toxicity of breast cancer treatments: from understanding to enhancing survivorship care. ESMO Open 2025;10:105128.

29. Cardinale D, Colombo A, Bacchiani G et al. Early detection of anthracycline cardiotoxicity and improvement with heart failure therapy. Circulation 2015;131:1981–8.

30. Romond EH, Perez EA, Bryant J, et al. Trastuzumab plus adjuvant chemotherapy for operable HER2-positive breast cancer. N Engl J Med. 2005;353(16):1673–1684.

31. Gianni L, Eiermann W, Semiglazov V, et al. Neoadjuvant chemotherapy with trastuzumab followed by adjuvant trastuzumab versus neoadjuvant chemotherapy alone, in patients with HER2-positive locally advanced breast cancer (the NOAH trial): a randomised controlled superiority trial with a parallel HER2-negative cohort. Lancet. 2010;375(9712):377–384.

32. Ewer MS, Vooletich MT, Durand JB, et al. Reversibility of trastuzumab-related cardiotoxicity: new insights based on clinical course and response to medical treatment. J Clin Oncol. 2005;23(31):7820–7826.

33. Burstein HJ, Lacchetti C, Anderson H, et al. Adjuvant Endocrine Therapy for Women With Hormone Receptor-Positive Breast Cancer: ASCO Clinical Practice Guideline Focused Update. J Clin Oncol. 2019;37(5):423–438.

34. Henson KE, McGale P, Taylor C, Darby SC. Radiation-related mortality from heart disease and lung cancer more than 20 years after radiotherapy for breast cancer. Br J Cancer. 2013;108(1):179–182.

35. Darby SC, McGale P, Taylor CW, Peto R. Long-term mortality from heart disease and lung cancer after radiotherapy for early breast cancer: prospective cohort study of about 300,000 women in US SEER cancer registries. Lancet Oncol. 2005;6(8):557–565.

36. Saiki H, Petersen IA, Scott CG, et al. Risk of Heart Failure With Preserved Ejection Fraction in Older Women After Contemporary Radiotherapy for Breast Cancer. Circulation. 2017;135(15):1388–1396.

37. Darby SC, Ewertz M, McGale P, et al. Risk of ischemic heart disease in women after radiotherapy for breast cancer. N Engl J Med. 2013;368(11):987–998.

38. Zellars R, Bravo PE, Tryggestad E, et al. SPECT analysis of cardiac perfusion changes after whole-breast/chest wall radiation therapy with or without active breathing coordinator: results of a randomized phase 3 trial. Int J Radiat Oncol Biol Phys. 2014;88(4):778–785.

